# Multi-resolution characterization of the COVID-19 pandemic: A unified framework and open-source tool

**DOI:** 10.1101/2021.03.12.21253496

**Authors:** Andy Shi, Sheila M. Gaynor, Corbin Quick, Xihong Lin

**Author notes:** Correspondence should be addressed to X.L. These authors contributed equally to this work (Andy Shi, Sheila M. Gaynor).

## Abstract

Amidst the continuing spread of COVID-19, real-time data analysis and visualization remain critical to track the pandemic’s impact and inform policy making. Multiple metrics have been considered to evaluate the spread, infection, and mortality of infectious diseases. For example, numbers of new cases and deaths provide measures of absolute impact within a given population and time frame, while the effective reproduction rate provides a measure of the rate of spread. It is critical to evaluate multiple metrics concurrently, as they provide complementary insights into the impact and current state of the pandemic. We describe a unified framework for estimating and quantifying the uncertainty in the smoothed daily effective reproduction number, case rate, and death rate in a region using log-linear models. We apply this framework to characterize COVID-19 impact at multiple geographic resolutions, including by US county and state as well as by country, demonstrating the variation across resolutions and the need for harmonized efforts to control the pandemic. We provide an open-source online dashboard for real-time analysis and visualization of multiple key metrics, which are critical to evaluate the impact of COVID-19 and make informed policy decisions.

## Introduction

The SARS-CoV-2 virus initially spread through local transmission in the Wuhan province of China and rapidly developed into a global pandemic with over 109 million documented cases worldwide as of 15 February 2021^1^. Various epicenters have emerged over the course of the pandemic on multiple continents, significantly impacting countries in Europe such as Italy and the United Kingdom and states in the United States such as New York and Florida, at various points in time. Efforts have been made in previous studies to understand transmission and spread, both at the individual level using contact tracing and at the population level using government and health department data. Nonetheless, the disease has continued to spread and resurge in previously affected regions due to persistent circulation of variants and emergence of new variants, to the detriment of the healthcare system, economy, and general welfare. Vaccination efforts have been broadening in order to control the pandemic’s spread; however, significant efforts including non-pharmaceutical interventions are still needed to mitigate continued risks and achieve global herd immunity to COVID-19^2,3^. The continual impact and spread of COVID-19 thus requires continued monitoring and tracking to manage the response to and impact of the pandemic. In this work, we present an analytical framework and online dashboard providing multiple metrics for daily tracking of COVID-19 spread and impact.

Many reports and web-based tools, often implemented as interactive dashboards, have been developed to track absolute measures of COVID-19 impact ^4^. Multiple governmental bodies have implemented trackers to provide testing, case, death and hospitalization counts; research groups have also implemented tools, such as the Johns Hopkins University Center for Systems Science and Engineering COVID-19 Dashboard ^1^, which provides daily case and death counts at multiple geographic levels and is broadly accessed by the research and general community. In addition to counts, case and death rates valuably provide the case and death impact by accounting for population size and allow for comparison across regions differing in population size. These absolute measures indicate the present and cumulative state of the pandemic.

There are many metrics for assessing COVID-19 spread that are valuable in assessing impact and guiding containment efforts; thus, a single metric such as case rate should not be used in isolation. A key, dynamic measure of the pandemic’s spread is the effective reproduction number, *R*_*t*_. It is a time-dependent measure of how fast the pandemic is spreading and is defined as the average number of people who become infected from an infectious person. This metric on the transmission rate is therefore a relative measure on the multiplicative scale. The effective reproductive number has been estimated in pandemic-affected populations ^5,6,7^, with estimates ranging from as low as 0.3 when estimated after stringent centralized quarantine in Wuhan, China to over 5 when estimated around the period of disease introduction in Germany; *R*_*t*_ has been estimated as 2-2.5 in the United States. Existing and former tools incorporating *R*_*t*_ are limited to a small set of geographic levels. Such tools include Rt.live, which provided up-to-date estimation by US state based on a generative model ^8^, Epiforecasts, which provides estimation based on an adjusted EpiEstim model ^9^, and a site providing COVID-19 R estimation for California ^10^ using the Wallinga-Teunis method ^11^. These tools have provided location- and metric-specific insight but are hence limited to specific geographic regions and metrics, which do not completely capture the manifold impact of COVID-19.

It is essential to accurately track transmission in real time across geographic domains, nationally and globally, in order to monitor circulating and newly infected cases towards containing the pandemic. As demonstrated in the United States, where the pandemic had notable early impact on coastal states with increasing impact throughout the middle of the country into the fall and the winter, different trends may occur in different locations. This can be a result of multiple features, such as government policies, seasonal weather, and demographic aspects. It is thus valuable to have unified approaches to estimating metrics across locales.

As the pandemic continues to have significant and renewed impact across locales amidst changing non-pharmaceutical intervention measures and increasing vaccination rollout, accurate, real-time metrics for assessing the evolving spread are critical. We developed the Visualizing COVID-19’s Effective Reproduction Number (*R*_*t*_) website (http://metrics.covid19-analysis.org), which uses a daily data feed to apply a flexible log-linear model to estimate multiple epidemiological measures for assessing COVID-19 impact: the daily effective reproduction number, case rate, and death rate. We use aggregated case and death counts to characterize the spread of the COVID-19 pandemic at the county, state, and country levels. This tool provides daily reporting of estimated metrics in the form of an interactive dashboard implemented in R Shiny, with open-source code provided. In contrast to existing tools, our dashboard allows for the quantification, visualization, and comparison of a set of metrics on COVID-19 spread across multiple geographic resolutions. This tool is informative for regularly assessing COVID-19 spread, evaluating the impact of public health interventions and vaccination efforts introduced to reduce disease transmission, and motivating disease containment approaches.

## Results

The present analysis includes data from 1 March 2020 to 15 February 2021; at the end of this period there were a total of 109 million reported cases globally. We evaluate the effective reproductive rate, case rate, and death rates across the globe and within the United States over this period for which sufficient cases were observed to allow stable estimation (Methods).

### Reporting multiple metrics over time demonstrates impacts in trends

We used a log-linear regression model to estimate the effective reproductive rate, case rate, and death rate of SARS-CoV-2 (see statistical model, Methods), and report in visualizations including maps. To assess the pandemic’s spread and impact, we estimated metrics at county, state, and national levels. We note that estimates require sufficient case counts in order to report a stable value. Figure 1 shows seasonal snapshots corresponding to the effective reproductive rate on 1 April 2020, 15 July 2020, 1 November 2020, and 15 February 2021. This figure demonstrates the dynamic nature of *R*_*t*_, showing that rates of spread have fluctuated over time. We observe the trajectory of spread from Asia to Europe, Africa, and the Americas; this has been evidenced by the evolution of the disease tracked via genetic COVID-19 strains. These snapshots demonstrate that many continents were impacted in Spring 2020, with higher rates concentrating in Europe, Asia, and Africa at the country level in Summer 2020.

**Figure 1:**
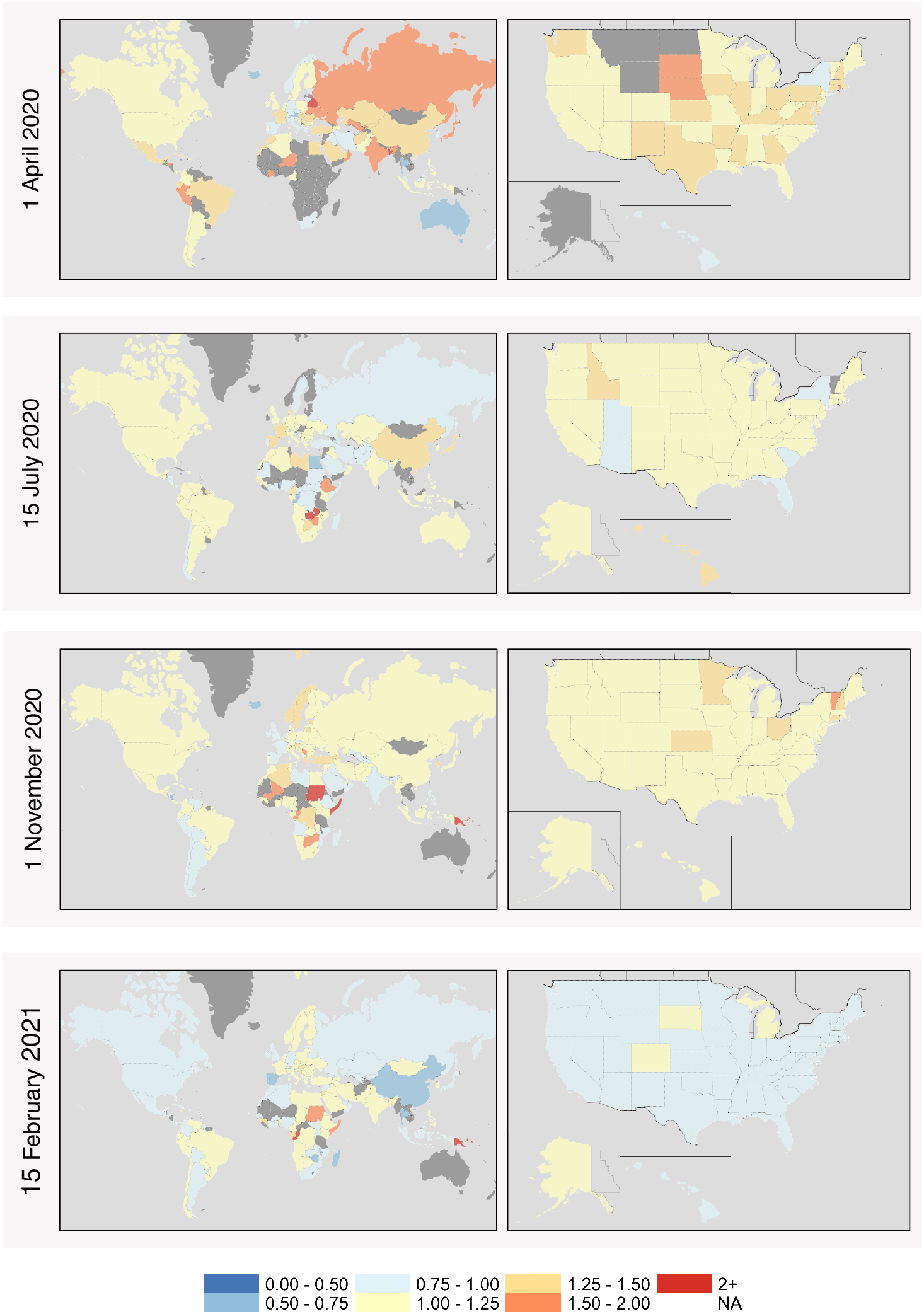
Effective reproductive number map over time. Global map of effective reproductive number *R*_*t*_ estimated for each country with sufficient data and US map of effective reproductive number *R*_*t*_ estimated for each state with sufficient data on 1 April 2020, 15 July 2020, 1 November 2020, and 15 February 2021, lagged by 7 days.

We further investigate the spread at two levels in the United States: states (Figure 1) and counties (Figure 4). In the US, states initially impacted in the spring demonstrated lower rates in Summer 2020 whereas Fall 2020 estimates indicate continued rates of spread across most states, decreasing in Winter 2021. Figure 2 further shows state-level measures, and demonstrates that there is a temporal ordering and complementary information provided by the three metrics. In particular, one can observe that in the Mid Atlantic region (New Jersey, New York, and Pennsylvania), *R*_*t*_ estimates primarily ranged from 1.00 to 1.50 from October through December 2020. This indicates that the disease was continuing to spread actively during this period, which corresponded to an increase in case rates continuously above 250 per million from November 2020 onwards, and a subsequent peak in death rates. Each of these metrics taken alone do not indicate the complete impact and potential for response; however, they collectively demonstrate that efforts during this period permitted epidemic-level spread which led to increased case and death rates.

**Figure 2:**
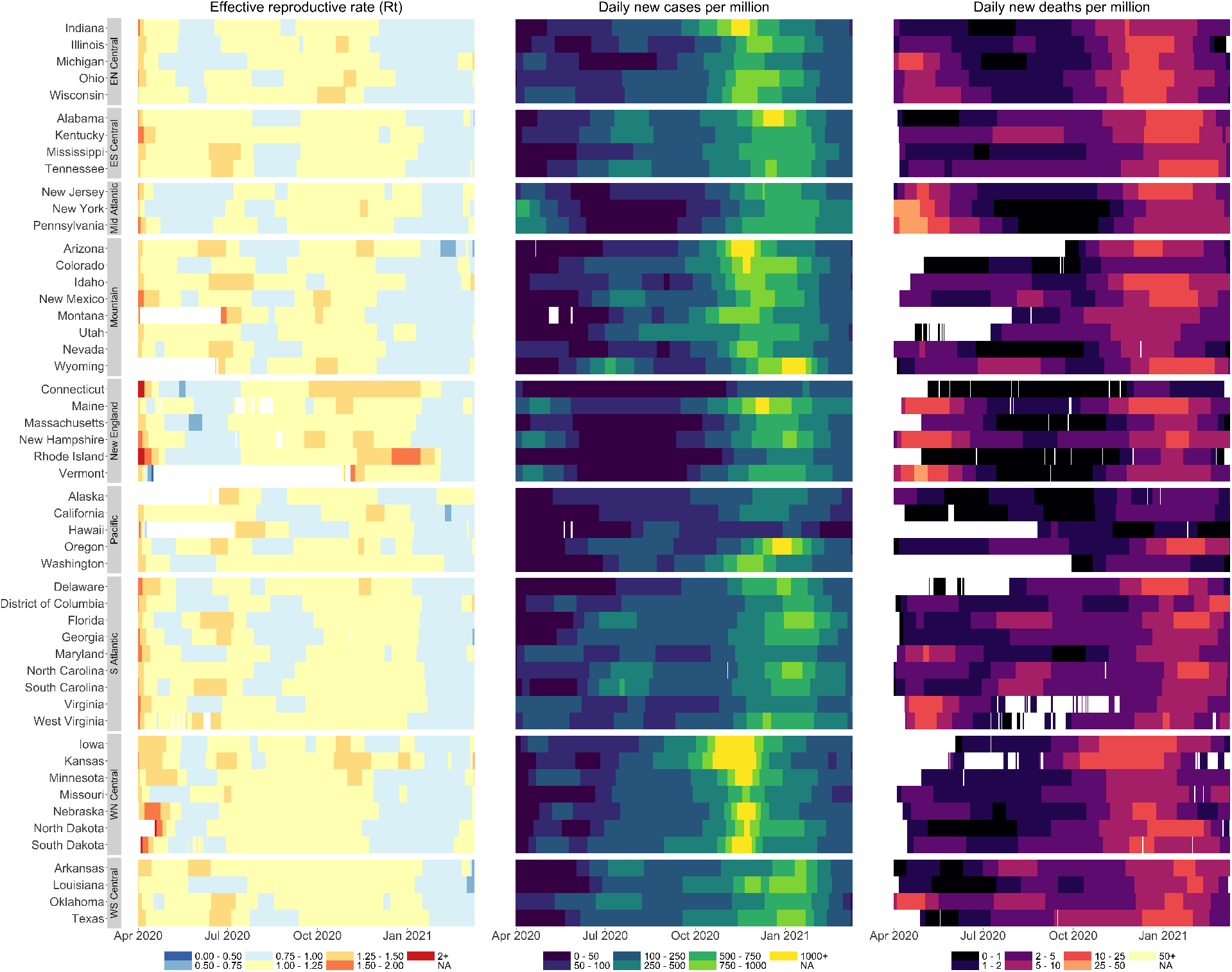
US State-level daily effective reproductive number, case rate, and death rate. Comparison of essential metrics *R*_*t*_ (left; lagged by 7 days), daily new cases per million (center), and daily new deaths per million (right) for each US state grouped by census region from 1 April 2020 to 15 February 2021.

**Figure 4:**
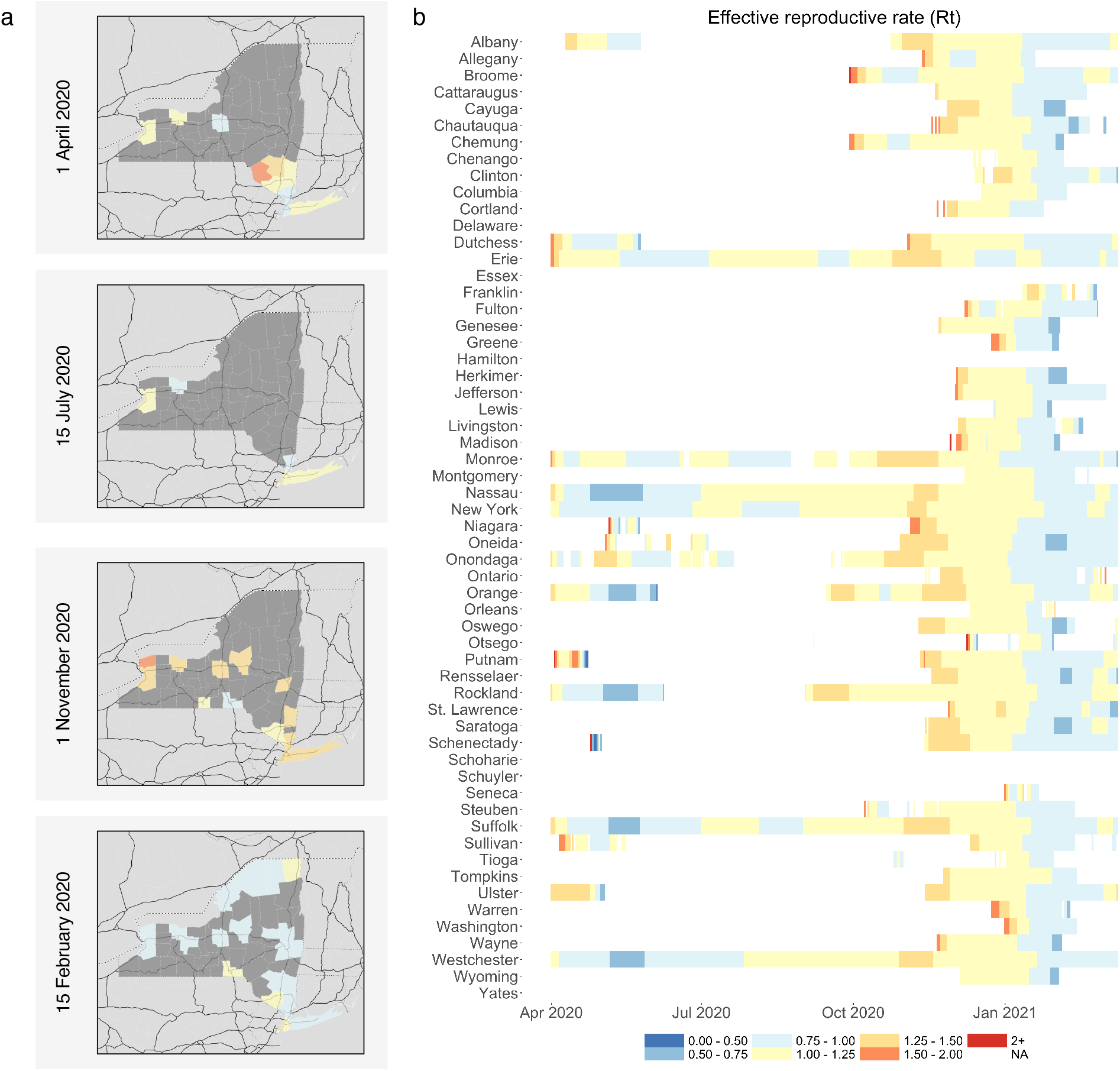
County-level effective reproductive number in New York. Comparison of effective reproductive number lagged by 7 days for (a) *R*_*t*_ mapped by NY county on 1 April 2020, 15 July 2020, and 1 November 2020, (b) *R*_*t*_ over time by NY county.

### Aggregate metrics can obscure trends at finer geographic resolutions

Many tools report metrics at a singular geographic resolution, such as at the country or state level. Given the population of 328 million in the United States and differences throughout the country in aspects such as demographics, health care access, population density, and pandemic response, it is understood that the pandemic’s impact has varied over time in the US. There is a significant amount of spatial variation in estimated metrics across geographic units, as demonstrated in Figures 1-3, where it is evident that state-level measures obscure more local trends in cases and spread. For instance, the rates at the US-level do not fully reflect regional differences at/within states. Figure 2 demonstrates state-level daily effect reproductive rate, case rate, and death rate grouped by census region. Regions such as New England with significant early impact are highlighted, while the Mountain and West North Central has sustained impact in later months. This suggests that the impact of the pandemic is heterogeneous across finer geographic levels, further suggesting that responses and interventions should also be targeted accordingly. Subnational units are provided for additional countries on the web resource.

In Figure 3a, the different trajectories of four US states are illustrated. While Florida demonstrated greater impact around the summer months, earlier impact in New York and later impact in South Dakota was clear. This illustrates that reporting at higher aggregate levels can obscure trends that can allow for more targeted interventions. At the county level, Figure 4 explores the trajectory of New York. The counties surrounding New York City experienced the most impact early on with *R*_*t*_ estimates regularly above 1.00, with improved containment state-wide in the summer of 2020. There was, however, rising spread in some upstate and western counties of New York in the fall of 2020, beyond that of the NYC region. This further illustrates heterogeneity in pandemic spread, even with a state. Given that strategies and resources are provisioned from multiple levels of governance and more generally that geographic units do not function in isolation, this demonstrates that accurate tracking requires study at multiple geographic resolutions.

**Figure 3:**
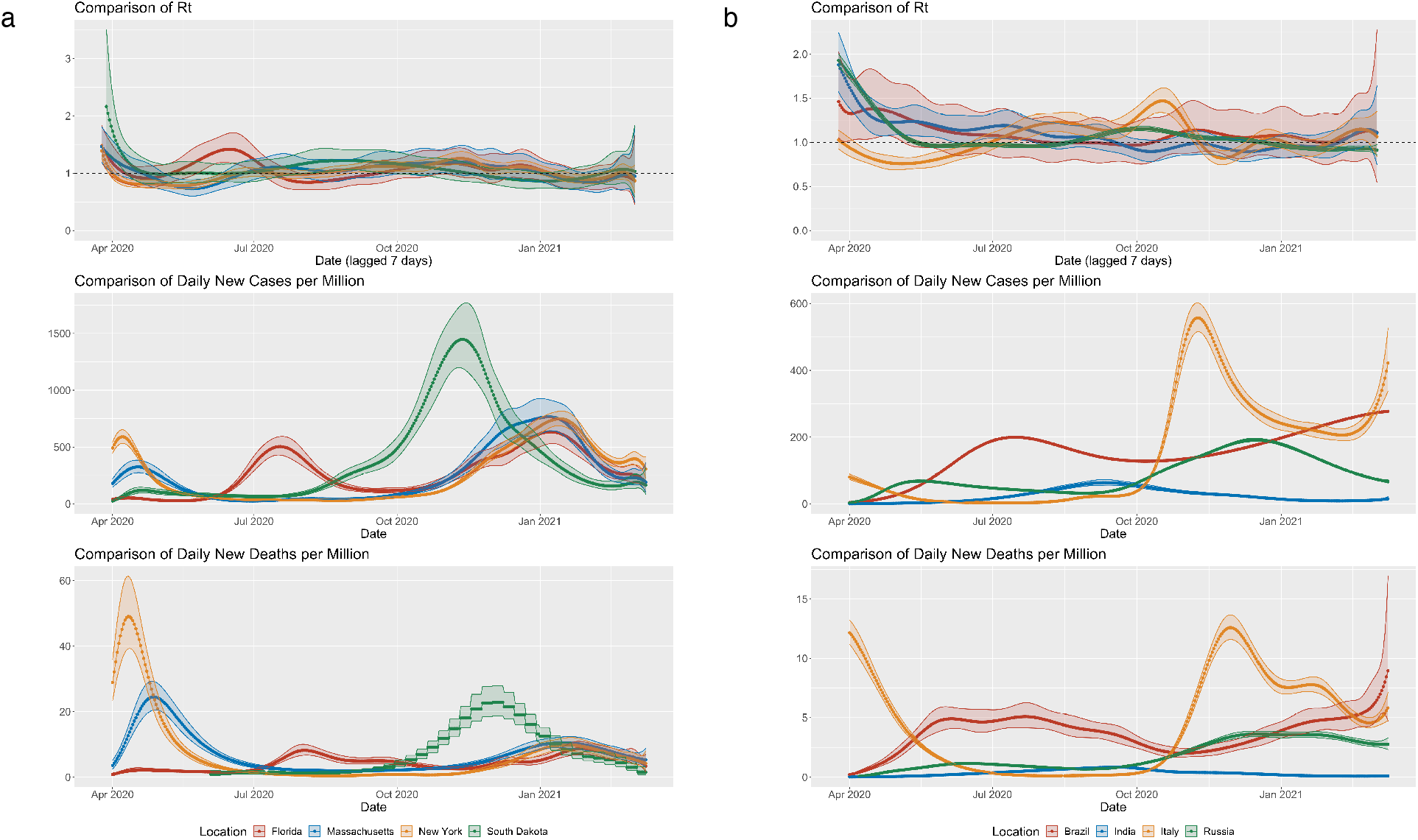
Country- and state-specific effective reproductive number, daily new cases per million, and daily new deaths per million over time. Comparison of essential metrics *R*_*t*_ (top panel), case rate (middle panel), and death rate (bottom panel) over time for US states and countries impacted by the pandemic. (a) Comparison of the US states of Florida (red), Massachusetts (blue), New York (orange), and South Dakota (green). (b) Comparison of the countries Brazil (red), India (blue), Italy (orange), and Russia (green).

### Daily reporting allows for real-time tracking

We leverage data from multiple reporting sources that have been aggregated and retrieved daily (see study data, Methods). To provide daily reporting, we evaluated metrics at multiple geographic resolutions from March 2020 to February 2021 in the present study, with real-time metrics available in the web resource. It is essential to provide regularly updated reporting in order to allow for corresponding action, while accounting for the fact that there can be case and death reporting delays and differences by weekday. Thus, we apply a framework that smoothly models the time-varying metrics from fitted estimates by using a B-spline for time and accounting for overdispersion on aggregated case and death data (see study data, Methods).

## Discussion

In this study, we perform a multi-resolution analysis of the transmission of SARS-CoV-2. We focus contributions on the estimation and evaluation of *R*_*t*_, as it is a key measure of disease spread and allows for tracking the pandemic trajectory. The use of cases and deaths alone do not fully characterize the pandemic’s spread for reasons such as difficulty in attributing deaths specifically to the virus, lack of reliability, or lag in counts. Thus, the effective reproduction number is an imperative measure to reduce in order to limit transmission and manage resources such as hospital beds ^12^.

There are significant implications for the reduction of *R*_*t*_ at across all levels. As demonstrated through the multi-level assessment of spread in the United States, evaluating spread at an aggregate state level can obscure trends occurring more locally, such as at the county level. While country and state level estimates can broadly characterize an area’s current impact, the reproduction rate is demonstrably heterogeneous. The importance of reduction is key given evidence of the impact of lack of access to care and over-extension of hospitals.

Many existing sites provide timely COVID-19 analyses; however, most are limited in scope to a particular analytic and geographic focus. Our tool critically provides a comprehensive and real-time suite of tracking and analyses at different geographical resolutions, including country, state and county levels. This is valuable as multiple metrics and considerations are necessary for providing a complete evaluation of the impact and status of the COVID-19 pandemic at each geographic resolution. Further, other tools oftentimes present data and analyses for specific geographic areas and thus are only relevant to particular communities. The software described here provides insights across multiple geographic levels globally.

Non-pharmaceutical interventions (NPI) have been implemented across societies to varying degrees in order to reduce transmission and control the spread of the pandemic; these measures have included social distancing, travel restrictions, and screening measures. Different interventions have varying effectiveness; similarly, currently approved vaccines have varying efficacy and are still being evaluated for their ability to block viral transmission ^3,13^. Timely reporting of *R*_*t*_ permits the evaluation of NPIs and vaccination efforts, and comparison across locales. Previous studies have analyzed the impact of various NPIs and systemic features in China and Europe ^6,14^, and recent studies have analyzed the impact of vaccination programs ^15^. The value of different sets of interventions and vaccination programs can be continually assessed using our unified framework.

There are multiple limitations of this work, including those attributable to the nature of a study performed on aggregated open data sources. Individual level data is not currently available to the public, limiting analyses to data that demonstrate features such as reporting lag over weekends due to testing and business operations and incomplete reporting due to governmental and clinical structure. Further, there are multiple potential modes of transmission, including symptomatic, presymptomic, asymptomatic and environmental transmission ^16^, which cannot be sufficiently studied without individual level data. A critical feature is that the analyses are limited to confirmed cases; there has been demonstrated limitations in access to testing and varying accuracy that has differed over time, which likely manifests as under-reported affected cases. The estimated reproductive numbers are valid if unascertaimnment rate does not change over time dramatically. Relatedly, there is documented spread from asymptomatic and subclinical cases that would not be captured by formal reporting through lack of testing.

Dashboards and web tools have provided a critical resource for the general public and policy makers during the pandemic. We provide multiple metrics to encourage the consideration of multiple, complementary measures to make informed decisions regarding interventions, behaviors and vaccination. The multiple levels and comparisons provided allow for viewers to target their regions of interest and understand the pandemic’s impact in their location. While there is an interplay of many factors contributing to the pandemic spread, trajectories of cases, deaths, and spread allow for understanding of and response to the pandemic’s impact on communities.

## Methods

### Study Data

We obtained daily confirmed COVID-19 cases and deaths at the county, state, and country level from the COVID-19 Data Repository by the Center for Systems Science and Engineering (CSSE) at Johns Hopkins University ^1^. This repository aggregated data primarily from government resources, such as Departments of Health, as well as news sources and other agencies. We retrieve data daily; for the present analyses data is as of 15 February 2021.

### Statistical Model

We use a generalized linear model (GLM) regression framework for estimating relevant metrics at various geographic levels, building upon the model by Cori et al ^17^. Let *Y*_*t*_ and *R*_*t*_ be the number of new cases and the effective reproduction number, respectively, on day *t* for a specific geographical location. As in Cori et al ^17^, we assume that, once infected, individuals have an infectivity profile given by a probability distribution *w*_*s*_, termed the serial interval, which depends on the time since infection of *s*. For example, an individual may be most infectious three days after becoming infected, so *w*_3_ would be greatest. The distribution of *w*_*s*_ depends on biological factors of the pathogen and is assumed to be known. In our analysis, we assumed *w*_*s*_ follows a discretized Gamma distribution with a mean of 5.2 days and standard deviation of 5.1 days ^18^. Then, let 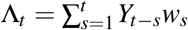 be the infectivity potential on day *t*. We assume that, conditional on the number of new cases on previous days, *Y*_*t*_ *∼* Pois(*R*_*t*_ Λ_*t*_). *R*_*t*_ is then the average number of secondary cases that each infected individual would infect assuming conditions remained the same at time *t*.

We extend this model by noting that log *E*(*Y*_*t*_ |Λ_*t*_) = log *R*_*t*_ + log Λ_*t*_. Therefore, we can consider modeling log *R*_*t*_ as a function of covariates and perform inference by fitting a Poisson model with log link to *Y*_*t*_, using log Λ_*t*_ as an offset. To smoothly model *R*_*t*_ as a function of time, we assume that log *R*_*t*_ = *B*(*t*)^T^*β*, where *B*(*t*)’s are the cubic B-spline basis and *β* is a vector of regression coefficients. We further extend this approach to account for overdispersion, and thus use a negative binomial model instead of Poisson. The maximum likelihood estimates and standard errors of *β* are obtained using Fisher scoring and then used to estimate *R*_*t*_ and calculate its confidence interval.

Because the number of reported cases on a particular day does not represent the number of people who contracted COVID-19 on that day, the *R*_*t*_ curve needs to be adjusted to account for the fact that people contract COVID-19 before their case is reported. We assume a 7-day lag from the time a person contracts COVID-19 until they are reported as a case, so we shift the *R*_*t*_ curve back 7 days to reflect this. This assumes an average incubation period of 7 days, which includes an average latent period of 3 days and an average presymptomatic period 2 days ^18^, plus an additional delay of two days to account for the time between getting tested and receiving a test result.

This model can then be extended to model the time-varying case rate or death rate. For case rate, we let *Y*_*t*_ be as before; for death rate, we let *Y*_*t*_ be the number of new deaths on day *t*. Then, the timevarying case/death rate is *ρ*_*t*_ = *Y*_*t*_ */C*_*t*_ where *C*_*t*_ is the population at time *t*. Therefore, we can again model log *E*(*Y*_*t*_ |*C*_*t*_) = log *ρ*_*t*_ + log*C*_*t*_ and model *ρ*_*t*_ using a cubic B-spline as log *ρ*_*t*_ = *B*(*t*)^T^*β*. We do not apply a lag to the case or death rate.

We further extended our model to handle several cases (see Supplementary Methods). This model flexibly permits time-varying estimation and it can further be used to evaluate the impact of time-varying non-pharmaceutical interventions. All R code for implementing this general model is publicly available at https://github.com/lin-lab/COVID19-Rt/.

## Data Availability

All materials and code for analysis are available on https://github.com/lin-lab/COVID19-Rt and https://github.com/lin-lab/COVID19-Viz.

https://github.com/lin-lab/COVID19-Viz

## Acknowledgements

This work was supported by a grant from the Partners in Health.

## Contributions

AS, SMG, and XL contributed to the conception of the work. AS, SMG, CQ, and XL contributed to the study design and method development. AS led the development of the online dashboard. AS and SMG contributed analysis and drafted the work. All authors revised the work and approved the submitted work.

