## Supplement for "Multi-resolution characterization of the COVID-19 pandemic: A unified framework and open-source tool"

### Supplementary Methods

We extended our log-linear model to handle certain edge cases.

#### Outlier Removal

First, we remove outliers from the data before fitting the models for $R_t$, case rate, and death rate. Outliers can occur when locations add a batch of historical cases that were previously unaccounted for. For example, on November 1, 2020, the US state of Georgia added about 30,000 historical cases based on antigen testing[1] and these cases were not back-distributed to when they actually occurred. Such outliers could greatly skew the model fitting, so we remove them when fitting the model. To remove outliers, we first compute the average number of new cases or new deaths in the past 7 weeks. Then, we remove days when the daily new count exceeded the average number of new cases in the past 7 days. However, if the number of such days is 10 or more, we do not remove any days' cases, because the data from this location is thus likely to be reported weekly. Next, we sort the daily new count (case or death) data in descending order. If the maximum daily new count is more than twice the next highest daily new count, then we subset the data to remove it. We repeat this process in the subset of the data until the largest daily new count data does not exceed twice the second largest.

Additionally, the number of daily new cases or deaths can sometimes be negative due to inaccuracies in tallying the cumulative number of cases. On those days, we impute a value of 0 new cases or new deaths.

#### Low Case or Death Counts

Next, we do not calculate $R_t$, case rate, or death rate when the number of daily new cases or deaths is small. On these days, the estimation procedure will produce wide confidence intervals due to the lack of data.

Therefore, we only fit calculate $R_t$, case rate, and death rate for a location after it has exceeded 50 total cases. Additionally, we do not calculate these metrics when the number of new cases is small. Specifically, we do not calculate $R_t$ when on days when there are fewer than 20 new cases on average in the past 7 days and we do not calculate the case or death rate when on days where there is fewer than 1 case on average in the past 7 days. This is justified given that our statistical model for $R_t$ assumes epidemic spread, which may not be occurring when the number of new cases is small. We also do not calculate $R_t$, case rate, or death rate for locations which have fewer than two weeks of data after removing outliers, removing dates with too few total cases, and removing dates where the 7-day average of new cases is too low.

#### Weekly Metrics

Furthermore, for certain locations we calculate a weekly $R_t$, case rate, or death rate. This is because certain locations may report data irregularly. For example, Nicaragua only releases COVID-19 data once per week. After removing outliers and dates where the total number of cases or 7-day average of new cases are below the thresholds in the previous section, we calculate a weekly $R_t$, case rate, or death rate if over 20% of the data are zero. The weekly case or death rates are then divided by seven to obtain an average daily case rate or death rate. For $R_t$, we set the weekly infectivity potential $\Lambda_t$ to be the number of cases in the previous week. In cases where the previous week had 0 new cases, we set $\Lambda_t$ to be the average of the new cases two weeks ago and average of the new cases during the current week.

#### Number of Knots for Splines

By default, we use a cubic B-spline with knots every 30 days. For the weekly $R_t$, case rate, and death rate, we use knots every 60 days instead because there are fewer data points. Additionally, when there is insufficient data, using knots every 30 days may lead to rank deficiencies. In those cases, we iteratively increase the knots by 15 days until there is no rank deficiency.

#### Overdispersion

The default negative binomial model assumed may not be appropriate where there is no overdispersion or when the variance function follows a different specification. The negative binomial has a variance function that is quadratic in the mean $\mu$: $V(\mu) = \mu + \mu^2/\theta$ where $\mu$ is the mean and $\theta$ is the inverse dispersion.

When there is no overdispersion, $\theta \to \infty$, which causes numerical issues during the model fitting step. In these cases or in other cases where the negative binomial model does not converge, we first fit a Poisson model. We then test for overdispersion by comparing the residual deviance to a $\chi^2$ distribution where the degrees of freedom are equal to the residual degrees of freedom. If the p-value obtained from this method is below 0.01, we fit a quasi-Poisson model instead, where the variance function is linear in the mean: $V(\mu) = \phi \mu$.

#### Additional Adjustments

For a small number (12 out of 3692) of localities, the default model parameters assumed are inappropriate and the estimated $R_t$, case rate, or death rate may have extremely high variability. After performing the main model fitting on all geographic localities, we examine the outputs for extremely large confidence intervals. For the small number of problematic localities, we adjust the outlier removal procedure, number of knots used for the B-spline or the cutoff for calculating weekly metrics. This post-hoc adjustment is needed because it would be unreasonable to assume the same model parameters hold for all locations worldwide.

### Sensitivity analysis

We performed sensitivity analyses to investigate sensitivity of effective reproduction rate estimates to the serial interval parameter estimates and the method for estimating the effective reproductive number. We first repeated the estimation of $R_t$ using our proposed approach while varying the serial interval based on a set of estimates in the literature as summarized in Appendix Table 1. The results of this sensitivity analysis given in Appendix Table 2 provides comparison via simple linear regression to the reported serial interval parameters of mean 5.2 and standard deviation 5.1. This analysis allowed the evaluation of different interval parameters derived from different populations, and demonstrated that estimates are largely consistent across serial interval estimates.

We next compared our approach to a different estimation method to evaluate the impact of modeling approach and assumptions on the estimation of the effective reproductive rate. We note that estimates are affected by errors and discrepancies in case and death reporting; we calculate estimates based on the provided data which has undergone previously described data checks. Systematic over or under reporting of counts could introduce bias into the estimates. The effect reproductive number was estimated using the EpiEstim approach[2], maintaining the same parameters as our method (SI Mean(SD): 5.2 (5.1)). The EpiEstim method imposes multiple assumptions, including a gamma prior distribution on Rt, constant Rt within a window given how highly variable Rt estimates are, and assumed time window width. Estimates are compared on dates when the total number of cases is at least 50 and the average number of new cases within the previous 7 days is at least 10 for stability. The methods comparison (Appendix Table 3) demonstrated that most estimates are moderately to highly correlated for these select dates.

Table 1: **Serial interval estimates in the literature.** Set of estimates of the serial interval mean and standard deviation as derived in different populations throughout the pandemic.

| Mean (SD) | Population | Reference |
|---|---|---|
| 3.96 (4.75) | Hubei, China | Du, Z., Xu, X., Wu, Y., Wang, L., Cowling, B. J., & Meyers, L. A. (2020). Serial interval of COVID-19 among publicly reported confirmed cases. Emerging infectious diseases, 26(6), 1341. |
| 4.4 (3.0) | Hong Kong | Zhao, S., Gao, D., Zhuang, Z., Chong, M. K., Cai, Y., Ran, J., ... & Wang, M. H. (2020). Estimating the serial interval of the novel coronavirus disease (COVID-19): A statistical analysis using the public data in Hong Kong from January 16 to February 15, 2020. medRxiv. |
| 4.7 (2.9) | Vietnam, South Korea, Germany, Taiwan, China, Singapore | Nishiura, H., Linton, N. M., & Akhmetzhanov, A. R. (2020). Serial interval of novel coronavirus (COVID-19) infections. International journal of infectious diseases, 93, 284-286. |
| 5.2 (5.1) | Wuhan, China | He, X., Lau, E. H., Wu, P., Deng, X., Wang, J., Hao, X., ... & Leung, G. M. (2020). Temporal dynamics in viral shedding and transmissibility of COVID-19. Nature medicine, 26(5), 672-675. |
| 7.5 (3.4) | Wuhan, China | Li, Q., Guan, X., Wu, P., Wang, X., Zhou, L., Tong, Y., ... & Feng, Z. (2020). Early transmission dynamics in Wuhan, China, of novel coronavirus–infected pneumonia. New England journal of medicine. |

Table 2: **Comparison of $R_t$ estimates across serial interval parameters.** Alternative serial interval parameters from the literature are compared to those used in the proposed method (SI Mean (SD): 5.2 (5.1)) for 1 April 2020, 15 July 2020, 1 November 2020, and 15 February 2021. Comparison is provided via simple linear regression (slope, intercept) for county, state, and country units.

| | SI Mean (SD) | 2020-04-01 | 2020-07-15 | 2020-11-01 | 2021-02-15 |
|---|---|---|---|---|---|
| County | 3.96 (4.75) | 1.016, 0.107 | 0.991, 0.016 | 0.943, 0.090 | 1.001, -0.042 |
| | 4.4 (3.0) | 0.996, 0.077 | 0.998, 0.023 | 1.042, -0.031 | 1.000, -0.025 |
| | 4.7 (2.9) | 0.923, 0.137 | 0.971, 0.048 | 1.023, -0.020 | 0.999, -0.018 |
| | 7.5 (3.4) | 0.320, 0.896 | 0.631, 0.386 | 0.691, 0.312 | 0.996, 0.046 |
| State | 3.96 (4.75) | 0.956, 0.180 | 1.191, -0.192 | 0.775, 0.278 | 0.678, 0.244 |
| | 4.4 (3.0) | 0.970, 0.106 | 1.091, -0.085 | 1.107, -0.105 | 1.122, -0.137 |
| | 4.7 (2.9) | 0.949, 0.086 | 1.039, -0.038 | 1.023, -0.020 | 1.027, -0.043 |
| | 7.5 (3.4) | 0.413, 0.676 | 0.630, 0.381 | 0.569, 0.462 | 0.837, 0.198 |
| Country | 3.96 (4.75) | 1.259, -0.233 | 1.004, -0.008 | 0.948, 0.058 | 1.006, -0.026 |
| | 4.4 (3.0) | 1.017, 0.026 | 1.016, -0.012 | 1.025, -0.013 | 1.020, -0.029 |
| | 4.7 (2.9) | 0.917, 0.123 | 0.956, 0.051 | 1.001, 0.009 | 1.011, -0.017 |
| | 7.5 (3.4) | 0.468, 0.526 | 0.605, 0.402 | 0.782, 0.214 | 0.924, 0.092 |

Table 3: **Comparison of $R_t$ estimates to existing method.** Alternative $R_t$ estimation using EpiEstim is compared to the proposed method, considering the same parameters (SI Mean (SD): 5.2 (5.1)) for 1 April 2020, 15 July 2020, 1 November 2020, and 15 February 2021. Comparison is provided via simple linear regression (slope, intercept) for county, state, and country units.

| | 2020-04-01 | 2020-07-15 | 2020-11-01 | 2021-02-15 |
|---|---|---|---|---|
| County | 0.798, 0.315 | 0.288, 0.758 | 0.562, 0.553 | 1.317, -0.319 |
| State | 0.612, 0.659 | 0.496, 0.557 | 0.552, 0.582 | 0.267, 0.632 |
| Country | 0.916, 0.150 | 0.199, 0.874 | 0.578, 0.444 | 0.673, 0.292 |
